# Preliminary Study on the Diagnostic Value of Ultrasonic Shear Wave Dispersion for High-risk Esophageal Gastric Varices in Cirrhosis

**DOI:** 10.1101/2022.11.11.22282074

**Authors:** Jiayin Wang, Hongyu Zhou, Tinghong Li, Chuan Liu, Lei Zhao, Baiguo Xu, Weili Yin, Fang Wang, Jing Liang, Xiang Jing, Huiling Xiang

## Abstract

**Objective:** To explore the clinical value of two-dimensional ultrasonic shear wave elastography (SWE) and shear wave dispersion (SWD) in the diagnosis of high-risk esophageal gastric varices with cirrhosis.

**Methods:** A total of 123 people were included, which were divided into the high-risk esophageal gastric varices (high-risk EGV) group (n =60) and low-risk esophageal gastric varices (low-risk EGV) group (n =63). Both shear wave elastography (SWE) and SWD were adopted to examine each patient’s liver synchronously with the Doppler ultrasonic instrument.

**Results:** In total patients, the results of SWD and SWE of the high-risk EGV group were significantly higher than low-risk EGV group respectively. According to SWD examination, the area under the curve (AUC) of high-risk EGV was 0.709(95%CI:0.616-0.802), the optimal cutoff value was 14.35 m/s/kHz, the sensitivity and specificity was 81.7% and 57.1%, the AUC of high-risk EGV in patients with compensated cirrhosis was 0.786(95%CI:0.656-0.916), the optimal cutoff value was 14.35 m/s/kHz, the sensitivity and specificity was 86.4% and 61.1%, while the AUC of high-risk EGV in patients with decompensated cirrhosis was 0.637(95%CI: 0.494-0.780). According to SWE, the AUC of high-risk EGV in all patients, patients with compensated cirrhosis, and patients with decompensated cirrhosis was 0.606(95%CI: 0.506-0.706), 0.596(95%CI: 0.449-0.743), and 0.579(95%CI: 0.434-0.725), respectively, indicating limited diagnostic value.

**Conclusion:** SWD predicted the existence of high-risk EGV in patients with compensated cirrhosis non-invasively and provided a supplementary method to determine whether high-risk EGV exists or not in patients, while SWE had limited diagnostic value.

## Introduction

Esophageal gastric varices (EGV) are common complications in patients with cirrhosis and portal hypertension. Despite the remarkable achievements in diagnosis and treatment of variceal bleeding,the mortality rate remains 12–22%. ^1,2^, which has a significant impact on the survival time and quality of life of the patients. Therefore, early detection of high-risk EGV is significant for improving the prognosis of the patients. Several guidelines ^2,3^ recommended that patients with cirrhosis should undergo gastroscopy to screen the extent of varicosity and the risk of bleeding; however, factors including the invasive nature of gastroscopy, poor tolerance of patients, and cost have limited its clinical application. With the development of detection devices and technology, noninvasive monitoring and early detection of high-risk EGV have become a research hotspot in recent years. According to ultrasound, shear wave elastography (SWE) is related to tissue stiffness, which has clinical value for predicting the existence of esophageal varices in patients with cirrhosis but has limited diagnostic value for high-risk EGV ^4^. Some studies found that SWD is positively correlated with tissue viscosity and can reflect the degree of inflammation and necrosis of the tissue ^5^. Previous studies on SWD mainly focused on patients with nonalcoholic steatohepatitis ^6.7^, while only a few studies were carried out in the population with cirrhosis. Therefore, this study explored the feasibility of SWD in evaluating high-risk EGV in patients with cirrhosis and studied the value of SWE in this population.

## METHODS

### Patients

Data of outpatients and inpatients with different types of cirrhosis were collected from the Department of Hepatology and Gastroenterology of Tianjin Third Central Hospital from February 2020 to 2021. This was a cross-sectional diagnostic accuracy experimental study.

Criteria for Enrollment: Patients with cirrhosis who have undergone SWE and SWD examinations of the liver and electronic endoscopy simultaneously.

Criteria for Exclusion: a. Patients who received endoscopic therapy in the past; b. Patients who underwent splenectomy in the past; c. Patients who underwent operation of the transjugular intrahepatic portosystemic shunt (TIPS) in the past; d. Patients with a confirmed diagnosis of liver cancer. No patients were treated with beta-blockers at the time of assessment.

According to the 2019 Guidelines for the diagnosis and treatment of liver cirrhosis^8^, the diagnosis of compensated liver cirrhosis is based on imaging examinations such as Type-B ultrasonography or computer tomography (CT) scan shows the characteristics of cirrhosis or portal hypertension, such as splenomegaly and diameter of portal vein ≥ 1.3 cm. The enrolled patients in this study have Type-B-ultrasound or CT imaging support. Decompensated cirrhosis is defined as the occurrence of any one of the complications, such as gastrointestinal bleeding, ascites, and hepatic encephalopathy. This study was approved by the Ethics Committee of Tianjin Third Central Hospital (Document No. IRB2019-022-03). All patients signed informed consent before participation in the study. Trial registration number: NCT03990753

### Abdominal ultrasound examination

A sonographer with > 10 years of experience, blinded to the examinations and laboratory results of the patients, performed the routine abdominal ultrasound and two-dimensional SWE/SWD examinations on the patients on a color ultrasonic instrument (Aplio I800, Canon Medical Systems, Tochigi, Japan). The probe was 3.5 MHZ, working by convex vibration. The operational method of SWE/SWD: The subject was fasted for 6 h, lying on the back, with the right arm stretched to the maximum extent and placed on the top of the head. The probe was perpendicular to the liver surface, trying to find the optimal intercostal window in the right lobe of the liver, avoiding large blood vessels and biliary ducts. The subject was trained to gently hold breath. The ultrasound system showed two views. After cooling for 5 s, the Quad-view mode was set, including shear wave elastography, propagation map, B-mode, and shear wave dispersion. A 3 cm×3 cm sampling frame was placed 1 cm below the liver capsule, and a 1 cm circular measurement area (region of interest (ROI)) was placed in the sampling frame to obtain liver stiffness measured in kPa and the dispersion measured in m/s/kHz. All operations were performed according to Canon’s manual of operation. All above examinations were conducted five times, and the average value was used. Following the initial guidelines for the elastography of the liver ^9^, we defined reliable measures of SWE as interquartile/median < 30%, and excluded patients with unreliable SWE results from the final analysis. (Figure.1)

**Fig 1.**
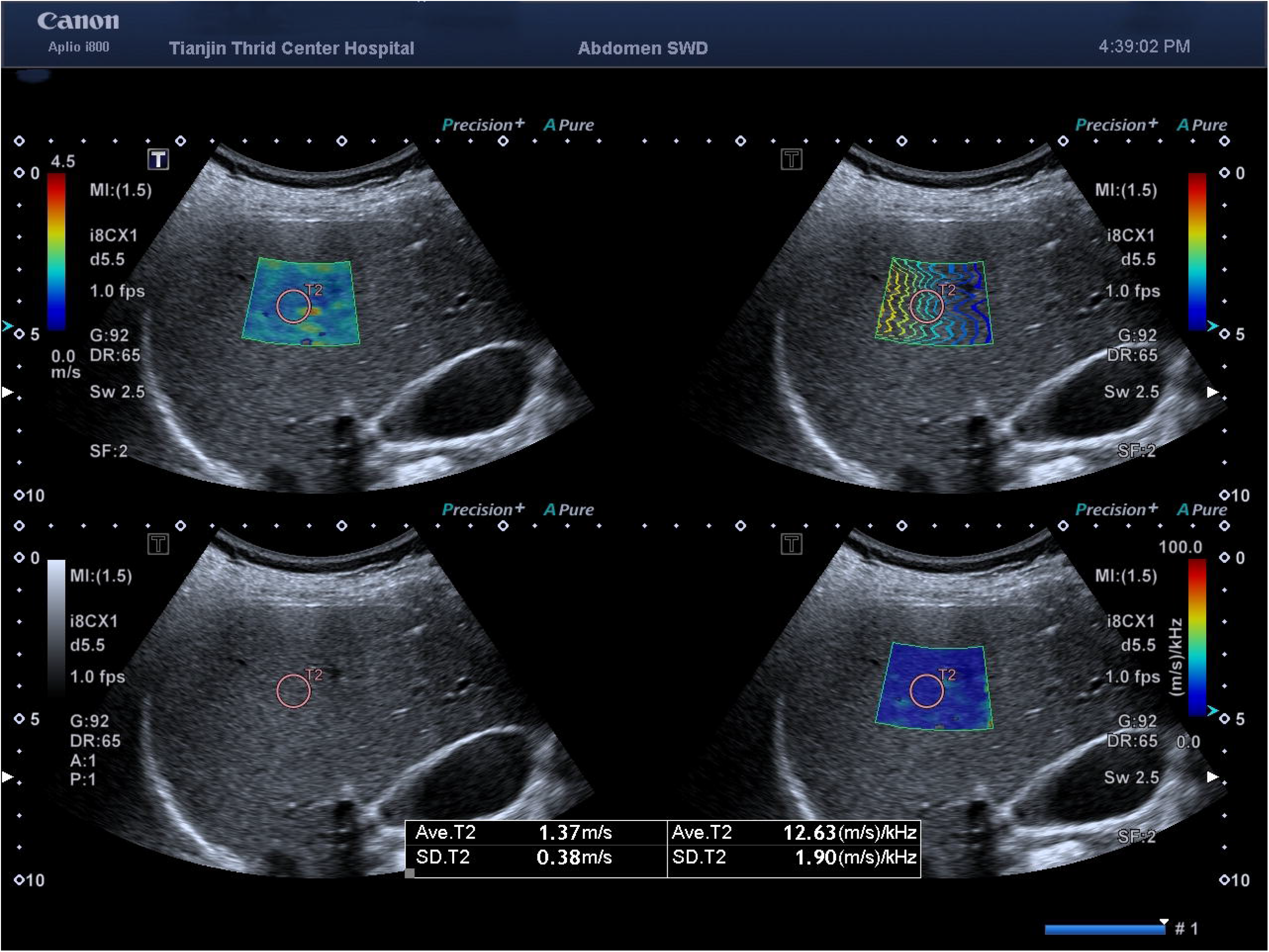
The Quad-view mode includes shear wave elastography (upper left), propagation map (upper right), B-mode (lower left), and shear wave dispersion (lower right).

### Endoscopy examination

All patients underwent a routine endoscopy examination (GIF-Q260J, Olympus, Tokyo, Japan) on an empty stomach. The results were checked by the same digestive physician with > 20 years of clinical experience but blinded to the other examination and test results of the patients. Parameters, such as the presence of EGV, and the location, diameter, and risk degree, were recorded according to the classification standard of location diameter risk factor (LDRf) ^2^

According to the standard of American association for the study of liver diseases (AASLD) ^10^, high-risk EGV is defined as medium/large varices (diameter ≥ 5 mm); small varices (< 5 mm) with red sign; small varices of decompensated cirrhosis.

### Blood test

Blood samples were withdrawn from the peripheral ulnar vein in fasting patients. The liver function test was carried out on Japan COBAS8000C-701 automatic biochemical analyzer. The liver function index assessed in this test was ALT (alanine aminotransferase).

### Statistical methods

Mean ± standard deviation was adopted for the measurement data conforming to the normal distribution, and t-test of two independent samples was adopted for comparison between the two groups; median and minimum, maximum [m (min, max)] were adopted for the measurement data conforming to nonnormal distribution. Mann–Whitney U test was adopted for comparison between the two groups; The classified variable was expressed as N (%). X^2^ test was adopted for comparison between the two groups. The receiver operating characteristic (ROC) curve was drawn to judge the diagnostic efficacy of SWD for high-risk EGV. P-value < 0.05 indicated a statistically significant difference. Statistical analysis was carried out using SPSS22.0 software.

## RESULTS

### Enrollment of subjects

In this study, the data of 235 patients with cirrhosis caused by various reasons were collected. The cohort included 171 patients who underwent endoscopy, SWD, and SWE at the same time, but excluded 37 patients who had received endoscopic treatment for varices, 9 patients who received splenectomy, 9 patients who had undergone the operation of TIPS, and 5 patients who had been diagnosed with liver cancer (some patients were overlapped). Finally, 123 patients with cirrhosis who fulfilled the inclusion criteria were enrolled in this study. According to the standard of AASLD, 63 patients were low-risk EGV and 60 patients were high-risk EGV. (Figure.2)

**Fig 2.**
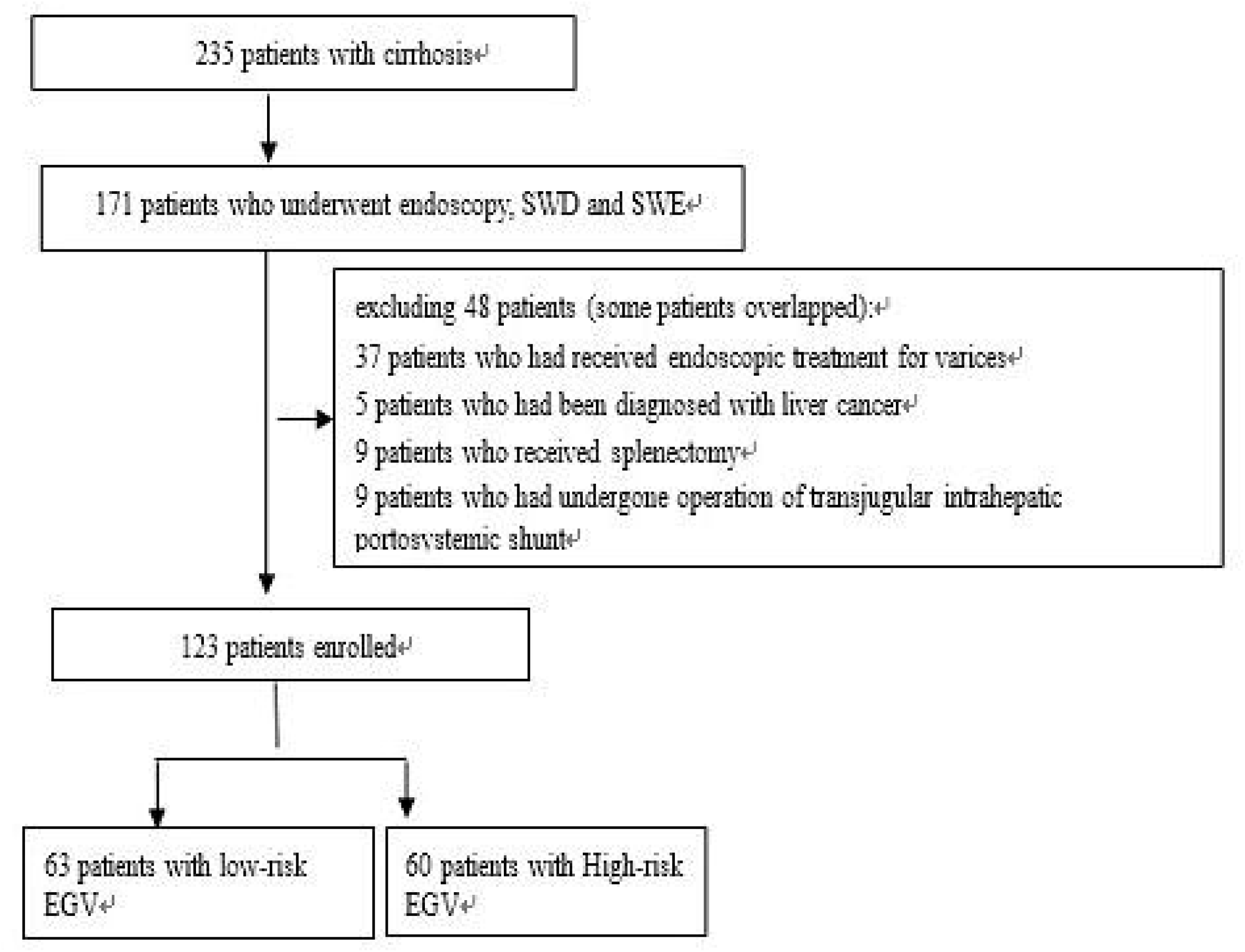
Flowchart of subjects

### General information of subjects

#### General information of patients in the compensated and decompensated patients

The average age of the subjects, including 72 males (58.5%) and 51 females (41.5%), recruited in this study was 55.12 ± 10.81 years old. Among them, 70 (56.9%) patients were divided into hepatitis B cirrhosis group, 11 (8.9%) into hepatitis C cirrhosis group, 18 (14.6%) into alcoholic liver disease group, and 24 (19.5%) into cirrhosis due to other causes group. 58 (47.2%) patients had compensated cirrhosis, and 65 (52.8%) had decompensated cirrhosis. Among the decompensated cirrhosis patients, 18 (27.69%) were divided into upper gastrointestinal bleeding, 52(80%) into ascites, and 3 (5.77%) into hepatic encephalopathy (some patients were overlapped). There was a difference in etiology between the two groups (P<0.05), and the other clinical baseline characteristics of both groups are presented in Table 1.

**Table.1.**
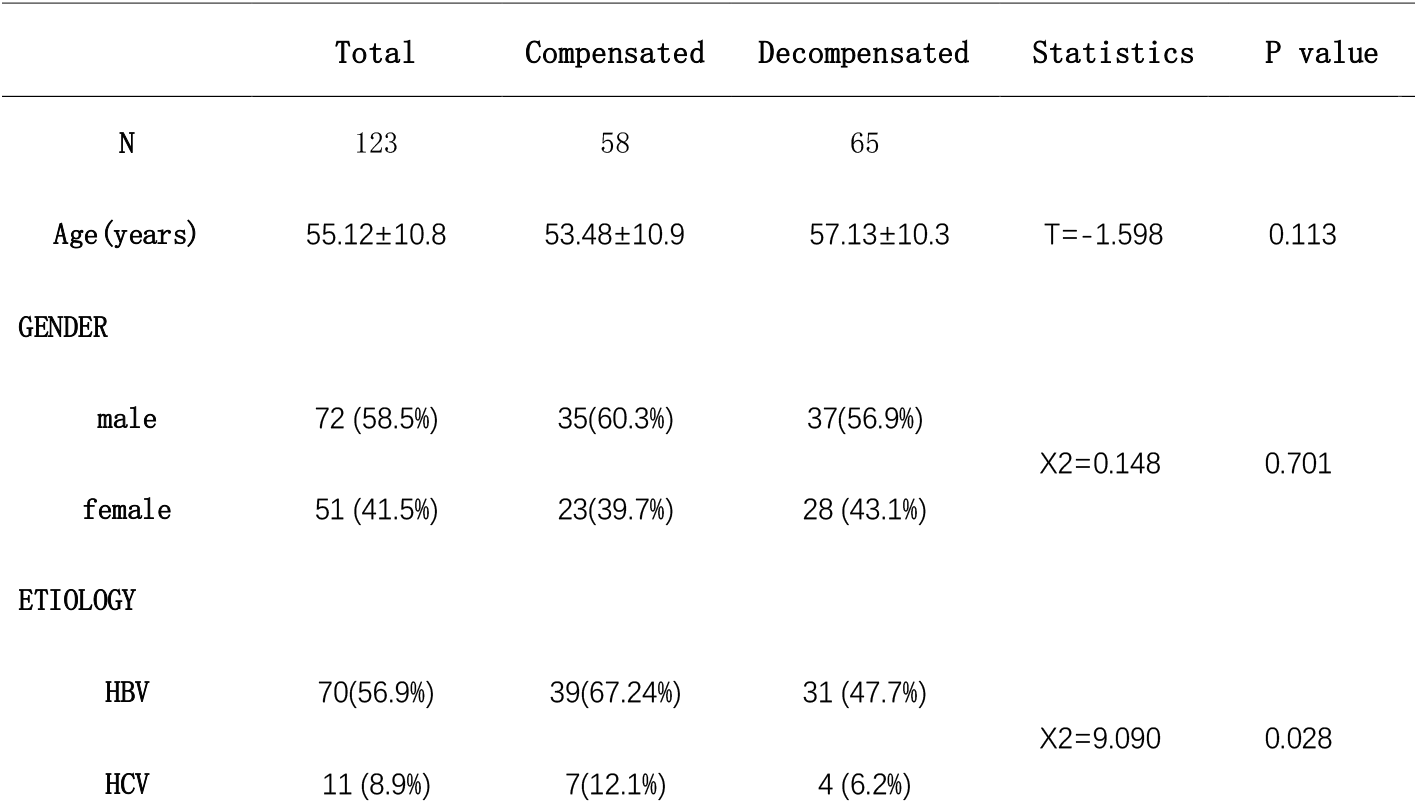
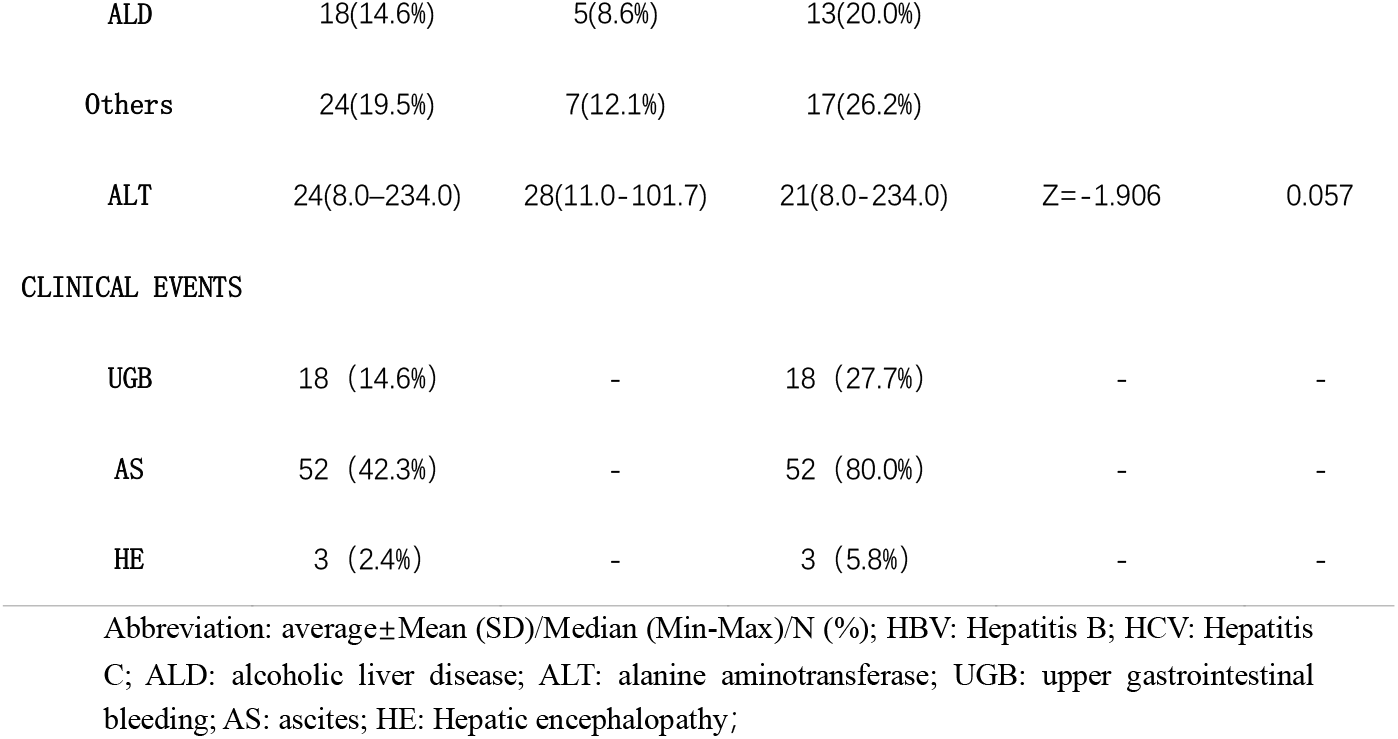
General information of patients with cirrhosis in the compensated and decompensated patients.

|  | Total | Compensated | Decompensated | Statistics | P value |
| --- | --- | --- | --- | --- | --- |
| N | 123 | 58 | 65 |  |  |
| Age (years) | 55.12±10.8 | 53.48±10.9 | 57.13±10.3 | T= -1.598 | 0.113 |
| GENDER |  |  |  |  |  |
| male | 72 (58.5%) | 35(60.3%) | 37(56.9%) | X2=0.148 | 0.701 |
| female | 51 (41.5%) | 23(39.7%) | 28 (43.1%) |  |  |
| ETIOLOGY |  |  |  |  |  |
| HBV | 70(56.9%) | 39(67.24%) | 31 (47.7%) | X2=9.090 | 0.028 |
| HCV | 11 (8.9%) | 7(12.1%) | 4 (6.2%) |  |  |
| ALD | 18(14.6%) | 5(8.6%) | 13(20.0%) |  |  |
| Others | 24(19.5%) | 7(12.1%) | 17(26.2%) |  |  |
| ALT | 24(8.0–234.0) | 28(11.0–101.7) | 21(8.0–234.0) | Z=-1.906 | 0.057 |
| CLINICAL EVENTS |  |  |  |  |  |
| UGB | 18 (14.6%) | - | 18 (27.7%) | - | - |
| AS | 52 (42.3%) | - | 52 (80.0%) | - | - |
| HE | 3 (2.4%) | - | 3 (5.8%) | - | - |

Abbreviation: average±Mean (SD)/Median (Min-Max)/N (%); HBV: Hepatitis B; HCV: Hepatitis C; ALD: alcoholic liver disease; ALT: alanine aminotransferase; UGB: upper gastrointestinal bleeding; AS: ascites; HE: Hepatic encephalopathy;

#### General information of patients with high-risk EGV and low-risk EGV groups

All patients were divided into 63 patients with high-risk EGV and 60 patients with low-risk EGV. There was no significant difference in age and gender between the two groups (P>0.05),which had significant differences in the etiology, ALT and whether decompensated or not. The other indicators of the two groups are shown in Table 2.

**Table.2.** General information of patients with cirrhosis in the high-risk EGV and low-risk EGV groups.

|  | Low-risk EGV | High-risk EGV | Statistics | P Value |
| --- | --- | --- | --- | --- |
|  | 63 | 60 |  |  |
| Age (years) | 55.2±10.5 | 55.0±11.2 | T=0.105 | 0.917 |
| Gender |  |  |  |  |
| male | (52.4%)(n=33) | (65.0%)(n=39) | X<sup>2</sup>=2.016 | 0.156 |
| female | (47.6%)(n=30) | (35.0%)(n=22) |  |  |
| Etiology |  |  |  |  |
| **HBV** | (68.3%)(n=43) | (45.0%)(n=27) |  |  |
| **HCV** | (11.1%)(n=7) | (6.7%)(n=4) | $\chi^2=13.077$ | 0.004 |
| **ALD** | (4.8%)(n=3) | (25.0%)(n=15) |  |  |
| **Others** | (15.9%)(n=10) | (23.3%)(n=14) |  |  |
| **Decompensated or not** |  |  |  |  |
| **no** | (57.1%)(n=36) | (36.7%)(n=22) | $\chi^2=5.171$ | 0.023 |
| **yes** | (42.9%)(n=27) | (63.3%)(n=38) |  |  |
| **ALT( U/L)** | 25.5 (8.0–234.0) | 23.0 (9.0–101.7) | $Z=-0.942$ | 0.346 |

Abbreviation: average ± Mean (SD)/Median (Min-Max); HBV: Hepatitis B; HCV: Hepatitis C; ALD: alcoholic liver disease; ALT: alanine aminotransferase;

### Distribution of SWD and SWE in the patients

In total patients, the mean of SWD of the high-risk EGV group was 16.43±2.75 m/s/kHz, which was significantly higher than 14.68±2.52 m/s/kHz of the low-risk EGV group (P < 0.001). In compensated patients, the mean of SWD of the high-risk EGV group was 16.59±2.66 m/s/kHz, which was significantly higher than 14.31±1.87 m/s/kHz of the low-risk EGV group (P < 0.001). There was no significant difference in patients with decompensated patients.

In total patients, the mean of SWE of the high-risk EGV group was 11.45 kPa, which was significantly higher than 9.4kPa of the low-risk EGV group (P =0.042). Compared with the high-risk EGV group and low-risk EGV group, there was no significant difference between the compensated patients and the decompensated patients. The specific values of both groups are presented in Table.3 and Figure.3.

**Table.3.** Distribution of SWD and SWE in the patients.

|  | Low-risk EGV | High-risk EGV | Statistics | P Value |
| --- | --- | --- | --- | --- |
| SWD(m/s/KHz) |  |  |  |  |
| Compensated | 14.3±1.0(n=36) | 16.6±2.7(n=22) | T=-3.844 | <0.001 |
| Decompensated | 15.2±3.17(n=27) | 16.3±2.8(n=38) | T=-1.543 | 0.128 |
| Total | 14.7±2.5(n=60) | 16.4±2.8(n=63) | T=-3.676 | <0.001 |
| SWE (KPa) |  |  |  |  |
| Compensated | 9.2 (4.7-34.2)<br>(n=36) | 10.45 (5.8-19.0)<br>(n=22) | Z=-1.218 | 0.223 |
| Decompensated | 10.5 (4.6-50.1)<br>(n=27) | 12.3 (5.5-28.0)<br>(n=38) | Z=-1.085 | 0.278 |
| Total | 9.4(4.6-50.1)<br>(n=60) | 11.5(5.5-28) (n=63) | Z=-2.029 | 0.042 |

Abbreviation: average±Mean (SD)/Median (Min-Max); SWD: shear wave dispersion; SWE: shear wave elastography;

**Figure 3.**
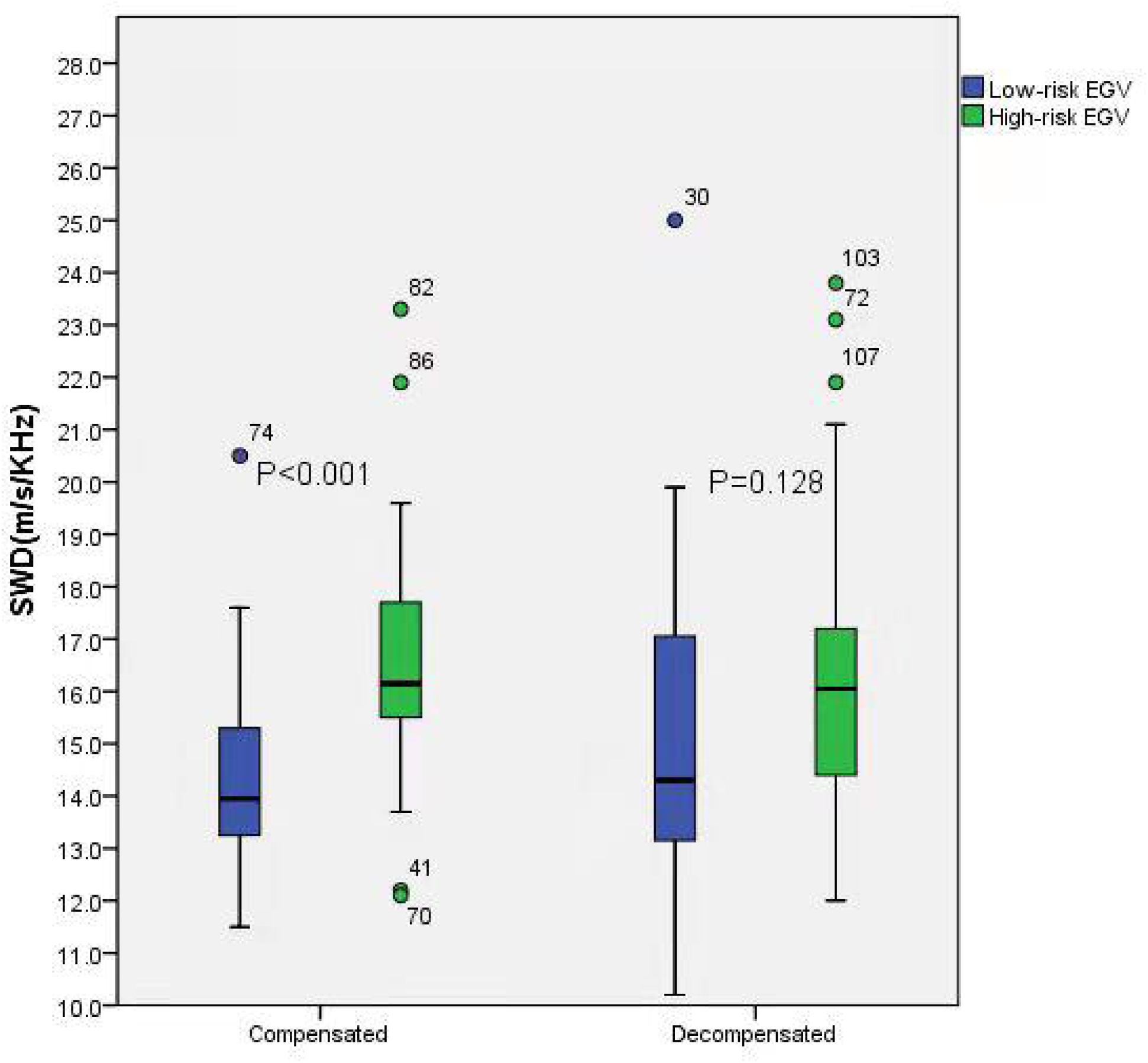
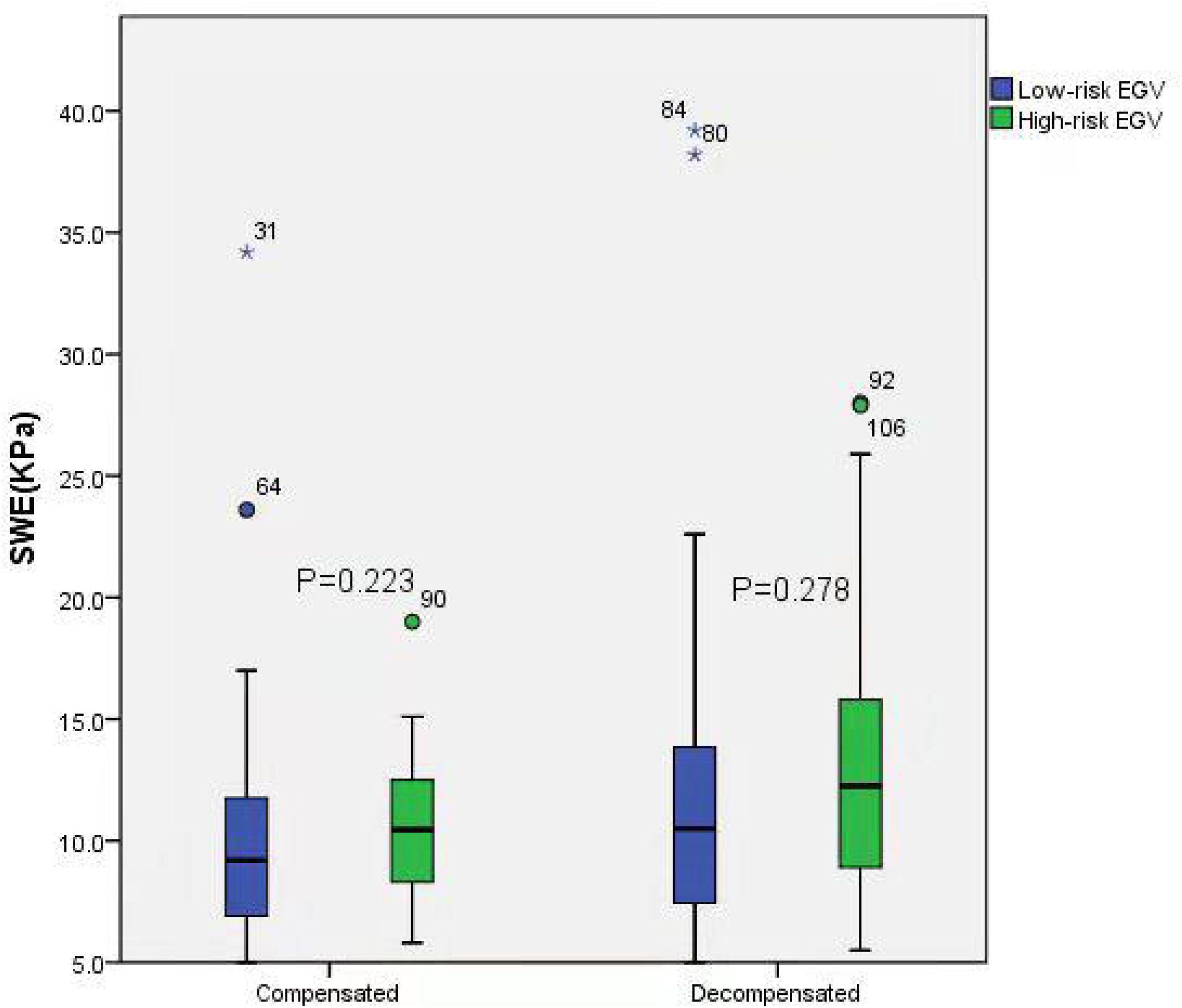
Correlation between SWD (A), SWE (B), and whether or not high-risk EGV in the compensated and decompensated groups. The box represents the quartile range (25th–75th percentile), and the middle line represents the median value of SWD/SWE. Significant differences in SWD between high-risk EGV and low-risk EGV in the compensated group, no significant difference was observed in SWE in the compensated and decompensated groups

### ROC analysis results

According to the diagnosis of SWD, the area under the curve (AUC) of total patients with high-risk EGV was 0.709, and the cutoff value calculated according to the Yorden index (sensitivity + specificity −1) was 15.35 m/s/kHz, with 68.3% sensitivity and 74.6% specificity. According to whether or not the patients were compensated, the AUC diagnosed by SWD in patients with compensated patients for high-risk EGV was increased to 0.786, while in decompensated patients, the AUC was 0.637. Moreover, the AUC diagnosed by SWE of patients with high-risk EGV in the total, compensated, and decompensated patients was 0.606, 0.596, and 0.579, respectively, which was of limited diagnostic value (Table.4 and Figure 4).

**Table 4.** ROC curve analysis of related indexes for diagnosing high-risk EGV in patients with cirrhosis.

| Test | AUROC | 95%CI | Cut-off | Specificity | Sensitivity | Accuracy | PPV | NPV |
| --- | --- | --- | --- | --- | --- | --- | --- | --- |
| **Subgroup analysis** |  |  |  |  |  |  |  |  |
| Compensated |  |  |  |  |  |  |  |  |
| SWE (KPa) | 0.596 | 0.449–0.743 | 9.25 | 52.8% | 68.2% | 60.5% | 59.1% | 62.4% |
| SWD (m/s/KHz) | 0.786 | 0.656–0.916 | 15.35 | 80.6% | 81.8% | 81.2% | 80.8% | 81.6% |
| Decompensated |  |  |  |  |  |  |  |  |
| SWE (KPa) | 0.579 | 0.434–0.725 | 14.7 | 81.5% | 36.8% | 59.2% | 66.5% | 56.3% |
| SWD (m/s/KHz) | 0.637 | 0.494–0.780 | 14.35 | 51.9% | 78.9% | 74.1% | 62.1% | 71.1% |
| **Total** |  |  |  |  |  |  |  |  |
| SWE (KPa) | 0.606 | 0.506–0.706 | 9.25 | 49.2% | 70% | 59.6% | 57.9% | 62.1% |
| SWD (m/s/KHz) | 0.709 | 0.616–0.802 | 15.35 | 74.6% | 68.3% | 71.5% | 72.9% | 70.2% |

Abbreviation: SWD: shear wave dispersion; SWE: shear wave elastography; PPV = positive predictive value; NPV = negative predictive value

**Figure 4.**
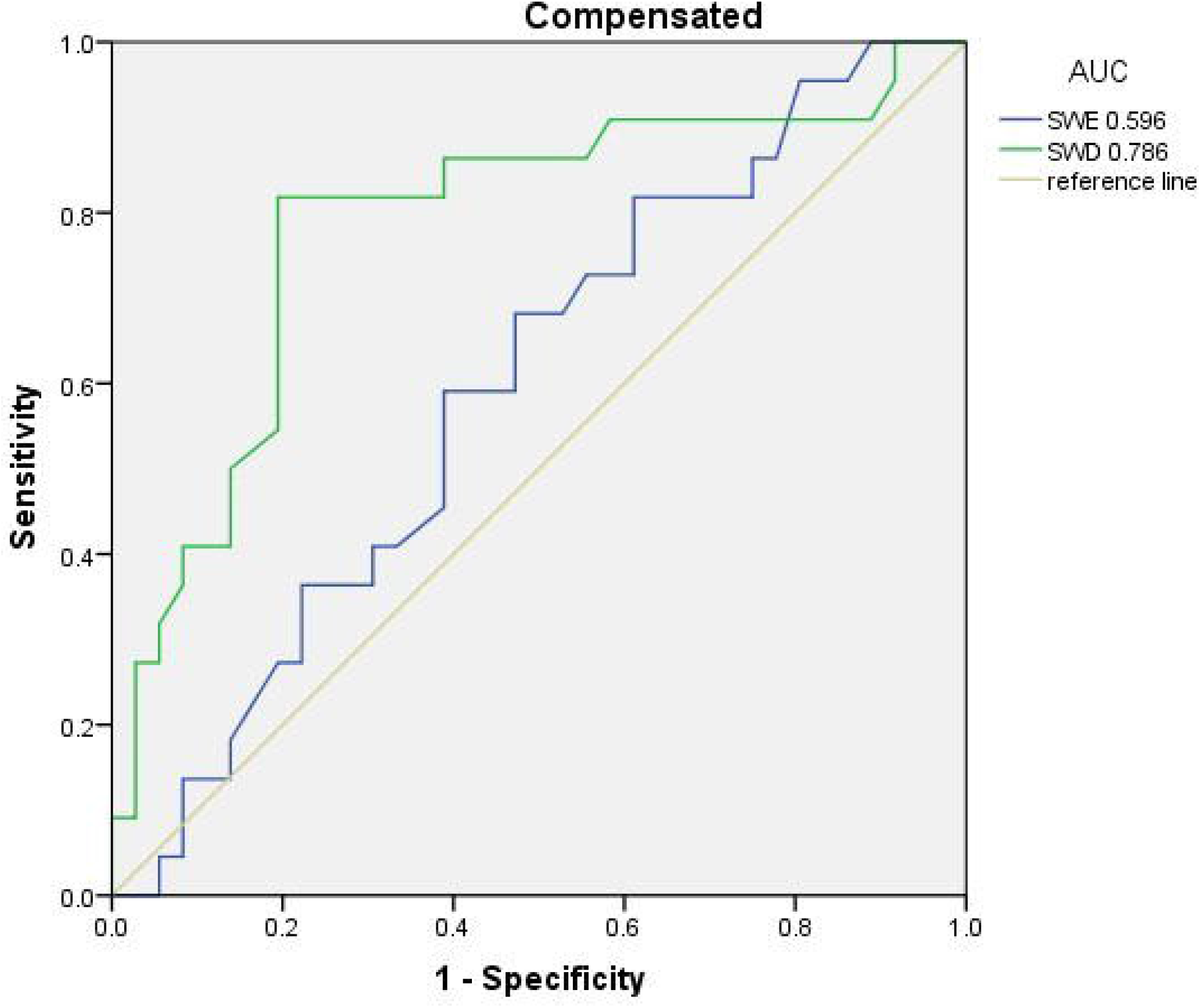
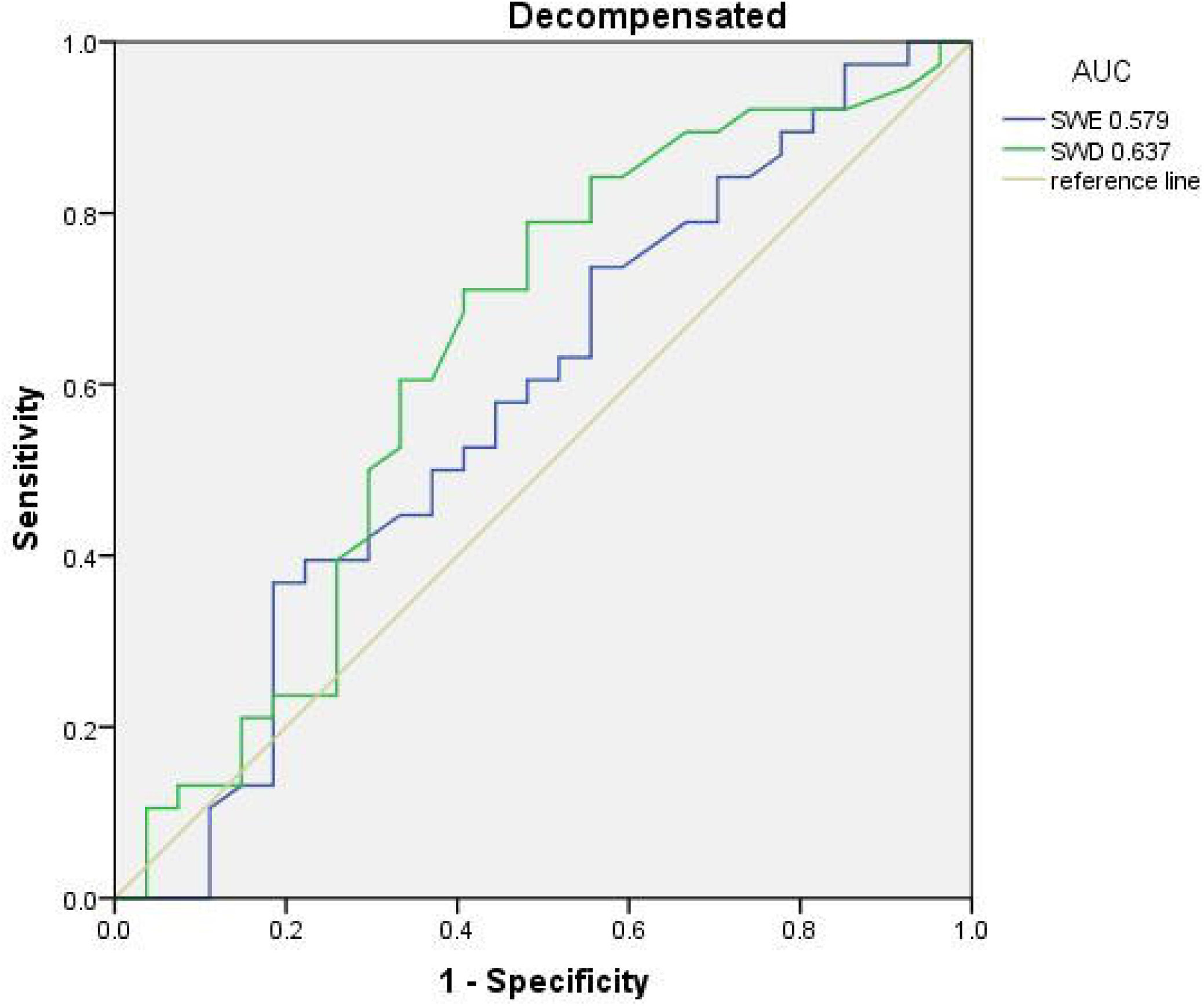
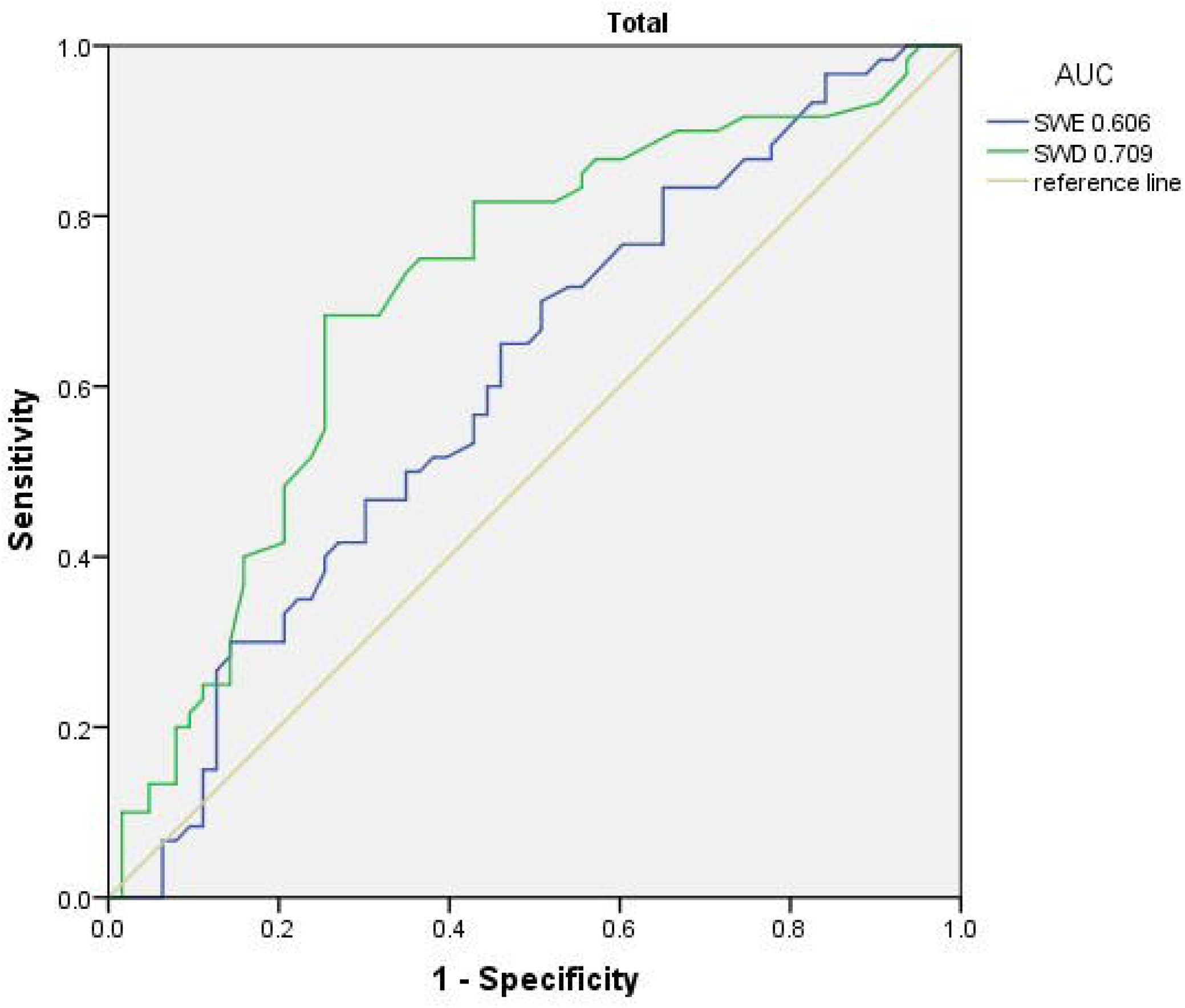
ROC curves of high-risk EGV in patients with cirrhosis diagnosed by SWD and SWE in the compensated group in figure (A), decompensated group in figure (B) and the total group in figure (C)

### Subgroup analysis of viral hepatitis and non-viral hepatitis

According to Table 4, we concluded that SWD had a high diagnostic value when predicting high-risk EGV in patients with compensated cirrhosis. Taking on account that etiology of liver disease affected the portal pressure by different way, Therefore, this study focused on the diagnostic value of the SWD in the two groups of the viral hepatitis and non-viral hepatitis. the AUC diagnosed by SWD in patients with compensated viral hepatitis patients for high-risk EGV was 0.777, with 84.6% sensitivity and 81.8% specificity, and in non-viral hepatitis patients, the AUC was 0.722. The specific values of both groups are presented in Table.5

**Table 5.**
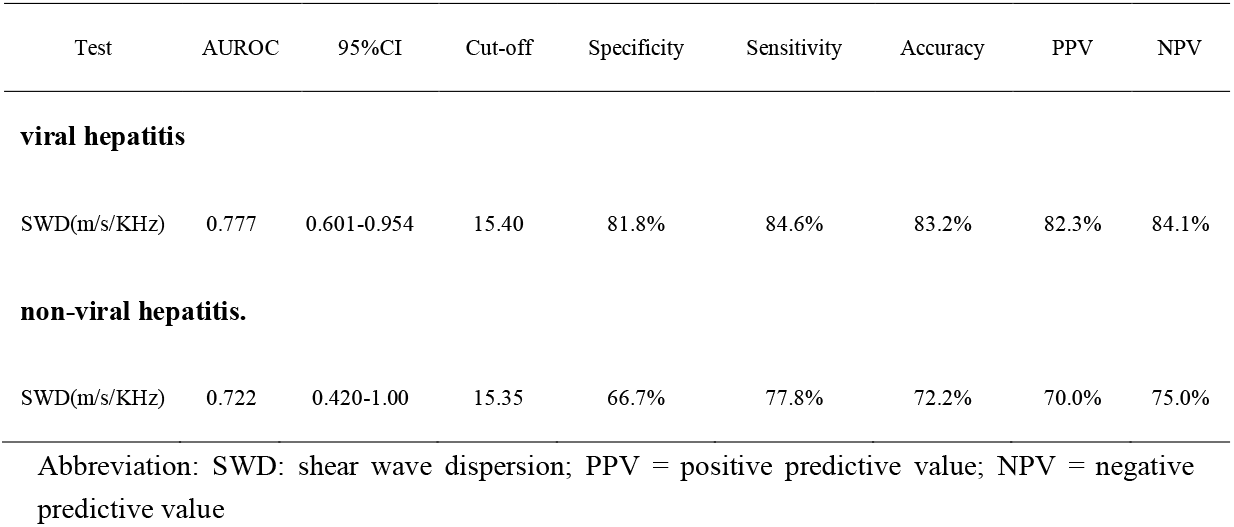
ROC curve analysis of related indexes for diagnosing high-risk EGV in the viral hepatitis group and non-viral hepatitis group.

| Test | AUROC | 95%CI | Cut-off | Specificity | Sensitivity | Accuracy | PPV | NPV |
| --- | --- | --- | --- | --- | --- | --- | --- | --- |
| **viral hepatitis** |  |  |  |  |  |  |  |  |
| SWD(m/s/KHz) | 0.777 | 0.601-0.954 | 15.40 | 81.8% | 84.6% | 83.2% | 82.3% | 84.1% |
| **non-viral hepatitis.** |  |  |  |  |  |  |  |  |
| SWD(m/s/KHz) | 0.722 | 0.420-1.00 | 15.35 | 66.7% | 77.8% | 72.2% | 70.0% | 75.0% |

Abbreviation: SWD: shear wave dispersion; PPV = positive predictive value; NPV = negative predictive value

## DISCUSSION

Cirrhosis is a disease caused by the long-term development of chronic liver diseases, such as extensive necrosis of liver cells, diffuse hyperplasia of liver fibrous tissues, and formation of false lobules or nodules. With the development of the disease, many complications may occur in patients with cirrhosis, including upper gastrointestinal bleeding that might be life-threatening^11^. Therefore, early detection of high-risk EGV is essential. The endoscopic examination exhibits the degree of varicosity, which is the gold standard for diagnosing EGV. However, because of its invasive and painful examination process, endoscopic examination is not accepted by patients, especially by those with asymptomatic cirrhosis, as it often affects the interventional treatment of patients. Therefore, the noninvasive assessment of varicosity in patients with cirrhosis has been emphasized.

In recent years, the study on the noninvasive diagnosis of EGV worldwide mainly focuses on PLT, ultrasound, magnetic resonance imaging (MRI), virtual hepatic vein pressure gradient and so on ^12-15^. However, these technologies are not yet perfect. For example, PLT indicators are easily affected by extrahepatic inflammation and clearance rate ^16^, and hence, it is necessary to combine other indicators (spleen diameter and liver stiffness) to improve the value of noninvasive indicators in predicting EGV. However, its accuracy in predicting EGV is limited, and AUC is between 0.71 and 0.84 ^17^. Magnetic resonance imaging (MRI), virtual hepatic vein pressure gradient, and other technical procedures are complex and expensive. Therefore, exploring a simple and practical noninvasive index is still the main research direction at present.

Ultrasonic elastography has gained increasing attention because of its low cost, high repeatability, and easy operation. Transient elastography (TE), alone or in combination with blood cell count and spleen size, could be used for noninvasive diagnosis of EGV in patients with cirrhosis^12^, which is encompassed by Baveno consensus; however, TE is affected by obesity, ascites, and intercostal space. Compared with TE, 2D-SWE integrates elastography into conventional ultrasound patterns, which showed the routine morphological structure of liver, blood flow, and liver stiffness of patients, and realized the “one-stop” evaluation of patient’s condition ^9^, which saved additional medical expenses. Moreover, the examination is visual, has a large sampling area, and is not affected by ascites, intercostal space, and obesity, which greatly improves the diagnostic accuracy and repeatability and is clinically applicable ^18^. SWE has a good diagnostic efficiency for esophageal varices, but its diagnostic value for the presence of high-risk EGV is limited ^4^. The main reason is that elastography assumes that the detected tissue is a pure elastomer without considering the effects of inflammation and necrosis. In a completely elastic tissue, the shear wave velocity is constant regardless of the shear wave frequency, while in viscoelastic tissue, the shear wave velocity changes with dispersion. Some studies have shown that the degree of lobular inflammatory activity is a critical determinant of SWD ^5, 19^. Cirrhosis is caused by an interactive process of inflammation, necrosis, and fibrosis. Therefore, SWE could not accurately reflect the real situation of the liver, and the factor of tissue viscosity should also be considered^20^. Thus, a new imaging technology software integrating SWE and SWD has been developed on Canon Aplio i series ultrasonic instrument. SWD is based on shear wave elastic imaging to further evaluate the tissue viscosity, which compensates for the deficiency of shear wave elastic imaging.

In this study, we found that the SWD of the high-risk EGV group in total patients was significantly higher than that of the low-risk EGV group (16.43±2.75 m/s/kHz vs. 14.68±2.52 m/s/kHz), and the AUC of high-risk EGV patients in SWD examination was 0.709. In patients with compensated patients,, the AUC was 0.786, while in decompensated patients, the AUC was 0.637.Taking on account that etiology of liver disease affects the portal pressure by different way, the AUC diagnosed by SWD in patients with compensated viral hepatitis patients for high-risk EGV was 0.777, with 84.6% sensitivity and 81.8% specificity, and in non-viral hepatitis patients, the AUC was 0.722. It could be seen that SWD had an advantage in patients with compensated cirrhosis patients, and compared to non-viral hepatitis patients, patients with viral hepatitis have higher diagnosis value… The two-dimensional shear wave dispersion (SWD) integrated conventional ultrasonic images together, Patients who completed SWD examinations could avoid repeated examinations of conventional ultrasound. What’s more, the measurement of SWD was simple, convenient, painless, reproducible, and easily acceptable. Before performing invasive endoscopy, it was expected to monitor SWD in real-time to avoid unnecessary endoscopy for patients with low-risk cirrhosis. The value of SWD in diagnosing high-risk EGV in patients with decompensated cirrhosis was limited, and AUC was 0.637. Recent studies ^21^ have shown that SWE may indicate portal pressure, but it cannot reflect the complicated hemodynamic changes of advanced portal hypertension. When the hepatic venous pressure gradient (HVPG) exceeds 10–12 mmHg, extrahepatic factors, such as vasoactive factors, spontaneous shunt channel formation, and portal vein thrombosis, have an increasing influence on portal pressure, while the proportion of liver factors are weakened, which is in line with the finding in this study that SWD has diagnostic value for compensated cirrhosis, but it is limited for decompensated cirrhosis.

In this study, the liver elasticity measured by SWE (AUC 0.694) is applicable in determining the presence of EGV, which will be presented in the future study of our center; however, its value is not high in diagnosing high-risk EGV. The AUC of high-risk EGV in SWE examination was 0.606 in the total group, 0.596 in the compensated group, and 0.579 in the decompensated cirrhosis group, which is inconsistent with previous studies ^22, 23^, and maybe related to different ultrasonic instruments, models, and versions ^24^. Moreover, SWE is affected by many factors. For example, the guidelines ^25^ pointed out that ALT would affect the diagnostic efficiency of SWE for fibrosis, and abnormal ALT in some subjects might affect the measurement of SWE. In comparison, SWD had considered the effect of inflammation, the result might be more stable than that of SWE.

Highlights of this study: The average value of SWD measured by Canon Aplio i800 in patients with cirrhosis was determined for the first time, and the clinical value of SWD in diagnosing high-risk EGV in patients with cirrhosis was discussed. Moreover, patients who completed SWD examinations could avoid repeated examinations of conventional ultrasound, reducing the cost of medical treatment for patients this study provided a supplementary mean for the noninvasive assessment of varicosity in patients with cirrhosis. It is worth expanding the sample for further discussion. Nevertheless, the following shortcomings cannot be ignored. First, the number of subjects was small. Second, the instrument used in this experiment was Canon ultrasonic series, which could not be extended to all machine models. Third, this study adopted the gold standard for endoscopy, and the evaluation of varice diameter was based on the doctor’s experience, lacking objective indicators. Fourth, the study recruited patients with cirrhosis, and the next step was to analyze the correlation between SWD and high-risk EGV.

Taken together, this study proves that the liver SWD slope of Canon Aplio i800 can provide a supplementary method for clinicians to predict the existence of high-risk EGV in patients with compensated cirrhosis, however, it is not sufficient to replace endoscopy.

## Data Availability

All data produced in the present study are available upon reasonable request to the authors

## Abbreviations

EGV: esophageal gastric varices
SWD: shear wave dispersion
SWE: shear wave elastography
TIPS: transjugular intrahepatic portosystemic shunt
LDRf: location diameter risk factor
AASLD: American association for the study of liver diseases
ALT: alanine aminotransferase
HBV: Hepatitis B
HCV: Hepatitis C
ALD: alcoholic liver disease
UGB: upper gastrointestinal bleeding
AS: ascites
HE: Hepatic encephalopathy
PPV: positive predictive value
NPV: negative predictive value
MRI: magnetic resonance imaging
TE: transient elastography
HVPG: hepatic venous pressure gradient

## Data Availability

The data used to support the findings of this study are available from the corresponding author upon request.

## Conflict of interest

The authors have no conflict of interests related to this publication.

## Authors’ Contributions

Jiayin Wang drafted this manuscript. Huiling Xiang. Hongyu Zhou, Tinghong Li, Lei Zhao, Baiguo Xu, Weili Yin, Fang Wang, Jing Liang, are responsible of the data collecting and management. Jiayin Wang, Huiling Xiang and Baiguo Xu are responsible of the data analyzing. Huiling Xiang, Xiang Jing, Chuan Liu designed the study and revised the manuscript.

Jiayin Wang and Hongyu Zhou contributed equally. The final version of the manuscript was reviewed and approved by all authors.

## Funding Statement

This study has received funding from the Tianjin Health Project. (No. TJWJ2022XK029)

### Trial registration number

NCT03990753

## Acknowledgements

Thanks to Professor Xiaolong Qi for his guidance on the research direction of the study. Thanks to Xinglin Chen for her guidance on statistical analysis.

## Supplementary Material

The STARD-Checklist of this study.

## Notes

### Competing Interest Statement

The authors have declared no competing interest.

### Funding Statement

Sponsored by Tianjin Health Project. (No. TJWJ2022XK029）

### Author Declarations

This study was approved by the Ethics Committee of Tianjin Third Central Hospital (Document No. IRB2019-022-03)

